# Mendelian randomization infers the effect of 14 parental illnesses on 44 congenital anomalies

**DOI:** 10.1101/2024.07.13.24310358

**Authors:** Tian Tian, Lei Tan, Wenjing Shao

## Abstract

**Background:** Congenital anomalies (CA), including congenital malformations (CM) and congenital deformities (CD), are significant health concerns influenced by genetic and environmental factors. Parental illnesses, especially those with genetic components, may affect the risk of congenital anomalies in offspring. Although clinical studies have suggested associations between certain parental illnesses and increased CM and CD risk, causal relationships remain unclear. This study employs a Mendelian randomization (MR) approach to investigate these potential causal links.

**Methods:** Fourteen parental illnesses were selected for this study: breast cancer, chronic bronchitis/emphysema, diabetes, heart disease, hypertension, and Alzheimer’s disease in mothers; and Alzheimer’s disease, bowel cancer, chronic bronchitis/emphysema, diabetes, heart disease, hypertension, lung cancer, and prostate cancer in fathers. Genetic variants associated with these illnesses were identified from genome-wide association studies (GWAS) in the UK Biobank. Genetic data for 44 congenital anomalies were sourced from the FinnGen database. Two-sample MR was conducted to estimate causal effects, with sensitivity analyses and multivariable MR (MVMR) to control for potential confounders.

**Results:** MR analysis revealed causal relationships between 13 parental illnesses and 13 specific congenital anomalies. Notably, mother’s hypertension significantly increased the risk of congenital hypothyroidism (IVW: OR = 7.969, 95% CI = 3.0826-20.6011, *p* = 4.20E-04), and father’s diabetes increased the risk of congenital heart defects in offspring (IVW: OR = 3.8E+09, 95% CI = 2.2E+04-6.6E+14, *p* = 3E-04). The associations’ strength varied with the type of parental illness and the specific congenital disease.

**Conclusion:** This study underscores the utility of MR in elucidating genetic influences of parental health conditions on congenital anomalies. The findings highlight the importance of managing parental health to reduce congenital anomalies risk in offspring. Further research is needed to explore underlying biological mechanisms and validate these findings in diverse populations.

## Introduction

Congenital anomalies(CA), encompassing a wide range of structural and functional anomalies present at birth, contribute significantly to infant morbidity and mortality worldwide[1]. Understanding the etiology of these conditions is crucial for developing preventative strategies and improving health outcomes. While environmental and genetic factors have been extensively studied, the influence of parental health conditions on the risk of congenital anomalies in offspring remains an area of ongoing research[2].

Parental illnesses, particularly chronic conditions, can have profound effects on the health and development of their children. These conditions may lead to adverse intrauterine environments, epigenetic modifications, or genetic predispositions that increase the risk of congenital anomalies. Observational studies have suggested associations between various parental illnesses and congenital anomalies. Recent studies have shown significant associations between parental chronic illnesses, such as diabetes, obesity, and hypertension, and the occurrence of congenital anomalies and other health issues in their offspring. For instance, maternal diabetes during pregnancy has been linked to congenital heart defects, neural tube defects, and other structural abnormalities in newborns[3–5]. Similarly, paternal health should not be overlooked. Research indicates that paternal obesity and metabolic syndrome are associated with an increased risk of cardiovascular diseases in children[6, 7].

Mendelian randomisation (MR) offers a robust approach to infer causality by using genetic variants as instrumental variables[8]. This method leverages the random assortment of genes from parents to offspring during meiosis, mimicking the randomised control trial design and thus reducing confounding[9]. By identifying genetic variants associated with specific parental illnesses, MR can provide insights into the causal relationships between these conditions and congenital anomalies.

In this study, we aim to investigate the causal effects of 14 parental illnesses on the incidence of 44 congenital anomalies in offspring using MR. By integrating genetic data from large-scale GWAS and relevant cohorts, we seek to elucidate the genetic underpinnings of congenital anomalies influenced by parental health conditions. Through rigorous MR analysis, we aspire to deliver reliable evidence that can guide future research and clinical practice in the prevention and management of congenital anomalies.

### Research Design and Methods

#### 1. Study Design and Data Sources

We employed both univariable Mendelian randomisation (UVMR) and multivariable Mendelian randomisation (MVMR) to investigate the causal effects of 14 parental illnesses on the incidence of 44 congenital anomalies in offspring. MR uses genetic variants as instrumental variables to infer causality, minimizing confounding factors and biases typically associated with observational studies. Throughout the study, we rigorously adhered to three core assumptions to ensure the validity of our results: 1. Establishing a reliable association between genetic variants and the risk factor. 2. Confirming no association between genetic variants and confounders. 3. Ensuring genetic variants solely influence the outcome through the risk factors[10]. A comprehensive study design flowchart is provided in Figure 1.

**Figure 1.**
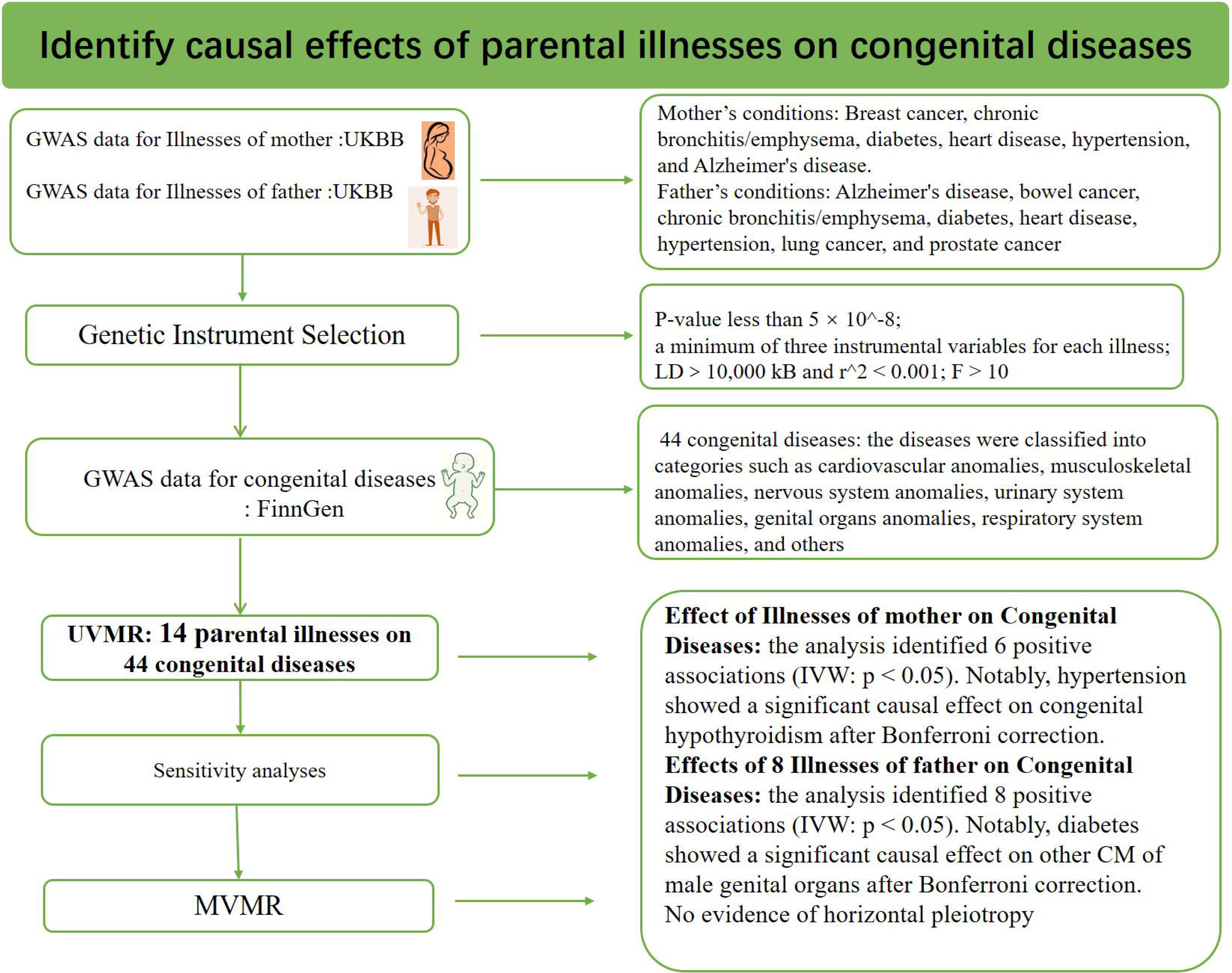
Flowchart of the study design. GWAS, genome-wide association studies; UVMR, Univariable MR; MVMR, multivariable mendelian randomization; UKBB, United Kingdom Biobank; IVs, instrument variables.

Our study utilized the largest and most recent publicly available summary statistics from multiple GWAS sources, including the FinnGen database and the UK Biobank (UKBB) database. To prevent overlap, we carefully selected data from the UKBB as exposure factors, and data from FinnGen were designated as the outcomes. Adherence to the STROBE-MR (Strengthening the Reporting of Mendelian Randomization Studies) guidelines was maintained throughout the study[11]. Detailed information on the GWAS data used in this study is provided in Supplementary Tables 1.

#### 2. Genetic Association Datasets

##### Parental Illnesses and Genetic Instruments

We sourced comprehensive GWAS data for parental illnesses from the largest study to date, selecting 14 parental illnesses from the UKBB. Each illness involved 300-400 thousand samples of European ancestry, encompassing approximately 10 million SNPs. The mother’s conditions included breast cancer, chronic bronchitis/emphysema, diabetes, heart disease, hypertension, and Alzheimer’s disease. The father’s conditions included Alzheimer’s disease, bowel cancer, chronic bronchitis/emphysema, diabetes, heart disease, hypertension, lung cancer, and prostate cancer. Detailed information on the GWAS data used in this study is provided in Supplementary Table 1.

##### Congenital anomalies

The study focused on 44 congenital anomalies, encompassing a wide range of structural and functional anomalies. Genetic data for these congenital anomalies were obtained from the FinnGen database, known for its comprehensive coverage of congenital anomalies[12]. Each disease involved 300 thousand samples of European ancestry and approximately 16 million SNPs. The diseases were classified into categories such as cardiovascular anomalies, musculoskeletal anomalies, nervous system anomalies, urinary system anomalies, genital organs anomalies, respiratory system anomalies, and others. Detailed information on the GWAS data used in this study is provided in Supplementary Table 1.

#### 3. Genetic Instrument Selection

For genetic instrument selection, we established a genome-wide significance threshold. SNPs with a P-value less than 5 × 10^-8 were considered significant. To ensure robust analyses, we required a minimum of three instrumental variables for each illness to perform the inverse-variance weighted (IVW) method and conduct sensitivity tests. Variants meeting these criteria were then clumped for linkage disequilibrium (LD) using a distance window of 10,000 kB and an r^2 < 0.001. To avoid the risk of weak instrumental bias, the F statistic was performed to evaluate the strength of the IV. When F > 10, the association between the IV and exposures was deemed to be sufficiently robust, thereby safeguarding the results of the MR analysis against potential weak instrumental bias. After several rounds of rigorous filtering, a set of eligible instrumental variables for the subsequent MR analysis were obtained. The instrument variables used in this study is presented in Supplementary Tables S5.

#### 4. Software

We employed the "TwoSampleMR”[13], "MendelianRandomization"[14], and "MR-PRESSO"[15] packages for UVMR and MVMR analyses, including sensitivity tests. Statistical analyses were conducted using R software version 4.3.2 (The R Foundation for Statistical Computing).

#### 5. Mendelian Randomization

##### Univariable MR Analysis

Causal effects were estimated using the random-effects inverse variance weighted (IVW) method. Causal estimates were expressed as odds ratios (ORs) with 95% confidence intervals (CIs). To ensure unbiased estimates, MR analyses were also conducted using four alternative methods: MR Egger, Simple mode, Weighted median, and Weighted mode. A causal effect was considered suggested if the IVW p-value was less than 0.05. Moreover, a causal effect was deemed significant if the IVW p-value fell below the Bonferroni-corrected threshold (p < 0.05/14 = 0.0035), coupled with consistent directionality in the weighted median and MR-Egger results.

##### Multivariable MR Analysis

For results from the UVMR analysis where multiple exposures were causally related to the same congenital disease, we performed MVMR to mitigate the confounding influence of the exposure factors. The MVMR-IVW method was used for this purpose[16].

##### Sensitivity analyses

Sensitivity analyses were conducted to validate the findings and to account for potential biases. MR-Egger Regression: Assesses directional pleiotropy and provides a pleiotropy-adjusted causal estimate. Weighted Median: Provides a robust causal estimate even if up to 50% of the information comes from invalid instruments[17]. MR-PRESSO: Detects and corrects for horizontal pleiotropy by identifying outlier SNPs[18]. Credible causal inference requires consistent directionality across the three methods and the absence of horizontal pleiotropic effects.

#### 6. Data and Resource Availability

All data generated or analyzed during this study are included in this article and its supplementary information files.

## Results

### Overview

The MR analysis investigated the causal effects of 14 parental illnesses on the incidence of 44 congenital anomalies in offspring. Among these, father’s bower cancer showed no causal effect on any of the 44 congenital anomalies. Thirteen other illnesses demonstrated causal relationships with 13 congenital anomalies (p < 0.05), with only two MR analysis results passing multiple testing. The remaining 31 congenital anomalies were not influenced by these 14 parental illnesses. The results highlighted significant associations between certain parental health conditions and specific congenital anomalies (Figure 2, Supplemental Table S2 and S3). Detailed results for notable associations are presented below.

**Figure 2.**
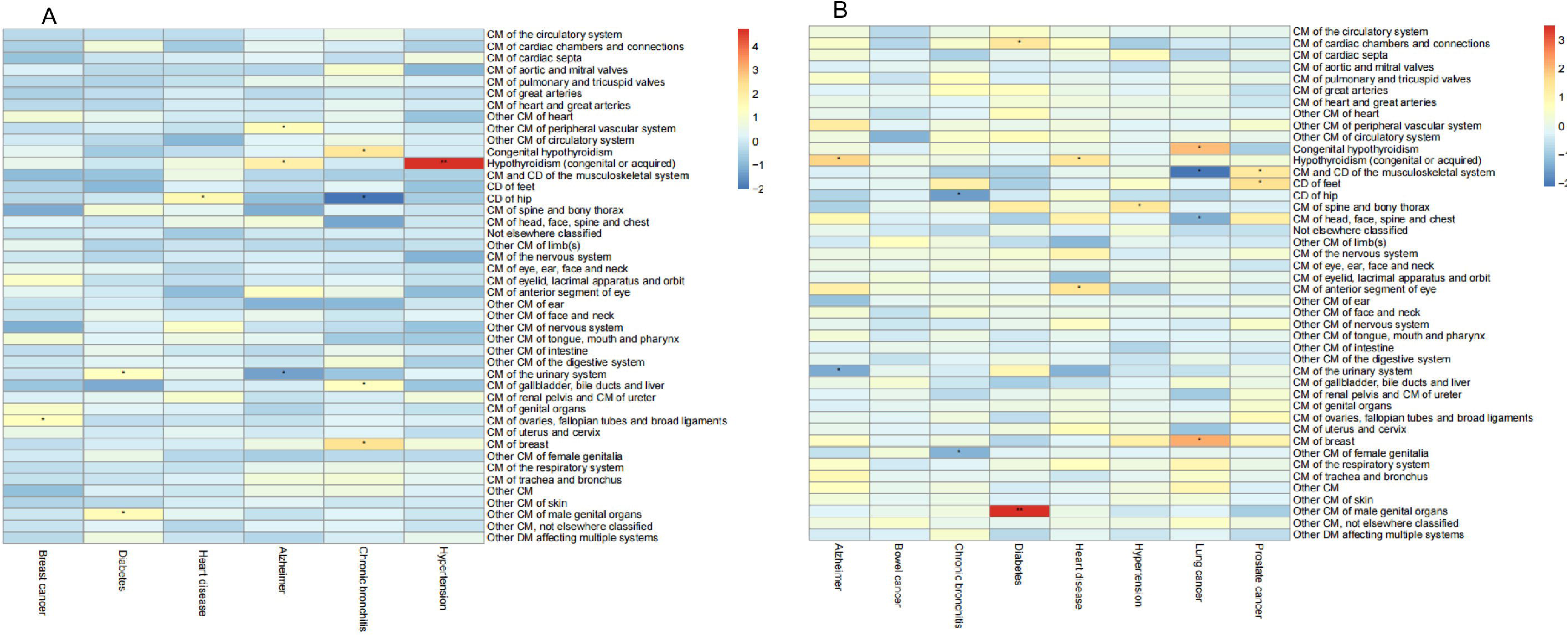
Heatmap of Mendelian randomisation effect size estimates (Signed Log-transformed p-value) of genetically predicted parental illnesses on congenital anomalies. (A) Effect of 6 illnesses of the mother on 44 congenital anomalies; (B) Effects of 8 illnesses of the father on 44 congenital anomalies. The Signed Log-transformed value represents the negative logarithm (base 10) of each p-value, multiplied by the sign of the effect. The asterisk ("*") indicates suggestive significance (p<0.05), while double asterisks ("**") denote significance after Bonferroni correction (p<0.0035 (0.05/14)). CA: Congenital anomalies; CD: Congenital deformities; CM: Congenital malformations.

### Effect of 6 Illnesses of mother on 44 Congenital anomalies

Using both UVMR(Figure 3) and MVMR(Figure 4), the analysis identified several positive associations (IVW: p < 0.05): breast cancer with congenital malformations (CM) of the ovaries, fallopian tubes, and broad ligaments; Alzheimer’s disease with other CM of the peripheral vascular system; diabetes with CM of the urinary system; hypertension with congenital hypothyroidism; chronic bronchitis/emphysema with CM of the breast; chronic bronchitis/emphysema with CM of the gallbladder, bile ducts, and liver; and heart disease with CD of the hip. Notably, hypertension showed a significant causal effect on congenital hypothyroidism after Bonferroni correction (IVW: OR = 7.969, 95% CI=3.0826-20.6011, *p* = 4.20E-04). (Figure 2, Supplemental Figure 2, Supplemental Tables 2). Sensitivity analyses revealed consistent results with no evidence of horizontal pleiotropy. One outlier was identified in the MR-PRESSO test for hypertension on congenital hypothyroidism; consistent results were obtained after removing the outlier (*p*=2.09E-06). Further MR analyses showed no causal links with other congenital anomalies. Additional results are detailed in Supplementary Table S2 and S4, with leave-one-out and scatter plots provided in Supplementary Figures 1-7.

**Figure 3:**
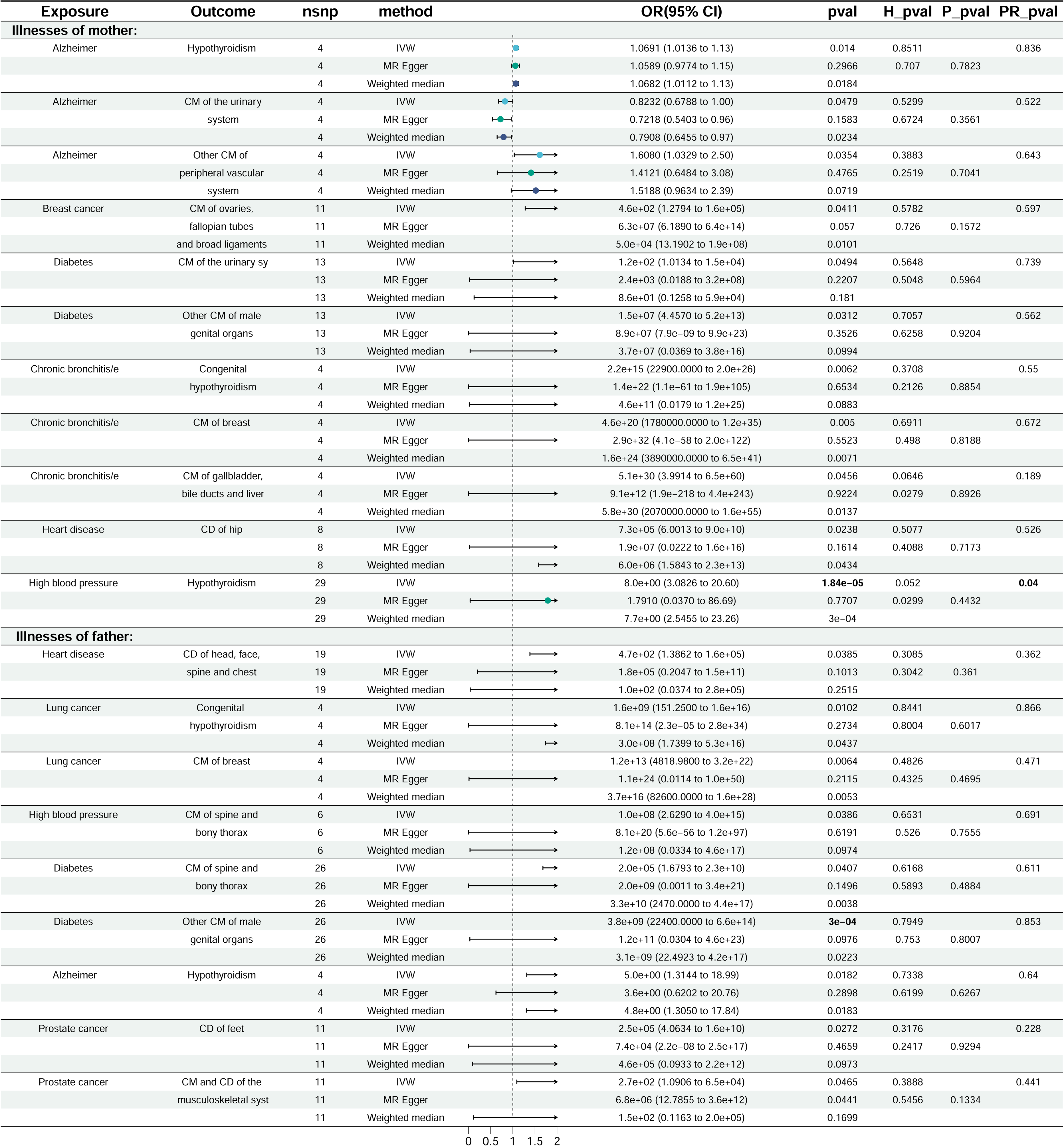
The UVMR of positive causal effects of parental illnesses on congenital anomalies. CA: Congenital anomalies; CD: Congenital deformities; CM: Congenital malformations; IVW: Inverse variance weighted; IVs: Instrumental variables; SNPs: Single Nucleotide Polymorphisms; MR: Mendelian randomization; UVMR: Univariable MR; OR: Odds ratio; CI: Confidence interval; GWAS: Genome-Wide Association Study; UKBB: United Kingdom Biobank. IVW p-values below the Bonferroni-corrected threshold (p<0.0035 (0.05/14)) are highlighted in boldface type.

**Figure 4:**
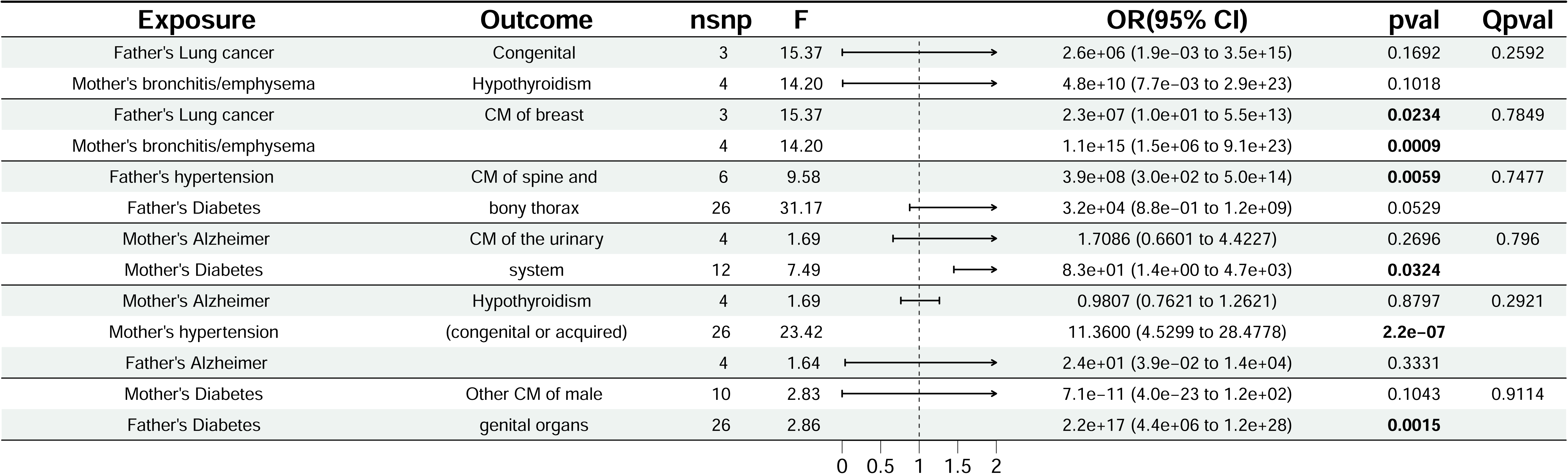
MVMR of parental illnesses on congenital anomalies. The MVMR-IVW method was used. Shown are the number of IVs in each analysis and the conditional F statistic for each exposure in MVMR. CA: Congenital anomalies; CD: Congenital deformities; CM: Congenital malformations; IVW: Inverse variance weighted; IVs: Instrumental variables; SNPs: Single Nucleotide Polymorphisms; MR: Mendelian randomization; MVMR: Multiple variable MR; OR: Odds ratio; CI: Confidence interval; GWAS: Genome-Wide Association Study; UKBB: United Kingdom Biobank.

### Effects of 8 Illnesses of father on 44 Congenital anomalies

Using instrumental variables for father’s illnesses, the following positive associations were found (IVW: p < 0.05) in UVMR (Figure 3) and MVMR (Figure 4): heart disease with CD of the head, face, spine, and chest; lung cancer with CM of the breast; hypertension with CM of the spine and bony thorax; diabetes with other CM of male genital organs; and prostate cancer with both CD of the feet and CM and CD of the musculoskeletal system. Most notably, after Bonferroni correction, the MR analysis for diabetes showed a significant causal effect on other CM of male genital organs (IVW: OR = 3.8 E+09, 95% CI=2.2 E+04-6.6 E+14, p = 3E-04). Sensitivity analyses confirmed no evidence of heterogeneity or horizontal pleiotropy. No outliers were detected by MR-PRESSO. Additional results are detailed in Supplementary Table S3 and S4. Leave-one-out analysis showed consistent results. Leave-one-out are provided in Supplementary Figures 8-13.

### MVMR

For results from the UVMR analysis where multiple exposures were causally related to the same congenital disease, we performed MVMR to mitigate the confounding influence of the exposure factors. The results revealed the following independent causal relationships: father’s lung cancer and mother’s bronchitis/emphysema with CM of the breast; father’s hypertension with CM of the spine and bony thorax; mother’s diabetes with CM of the urinary system; mother’s hypertension with congenital or acquired hypothyroidism; and father’s diabetes with other CM of male genital organs (Figure 4).

## Discussion

Our study provides novel insights into the causal relationships between parental illnesses and congenital anomalies in offspring through the application of Mendelian randomization. The significant associations observed, particularly the increased risk of congenital hypothyroidism associated with mother’s hypertension and CM of male genital organs associated with father’s diabetes, underscore the potential for parental health to influence fetal development. These findings enhance our understanding of the differing roles of parental health conditions in the development of congenital anomalies and provide avenues for further mechanistic investigation. However, our MR approach strengthens these associations by reducing the likelihood of confounding factors and reverse causation.

Our analysis reveals an unexpected causal relationship between mother’s hypertension and congenital hypothyroidism, suggesting a more intricate connection between mother’s health and offspring endocrine development than previously recognized. This association prompts a reexamination of the factors influencing the fetal thyroid gland’s maturation and function, potentially implicating a range of mechanisms from direct hormonal impacts to the influence of the in-utero environment on gene expression and developmental programming. Mother’s hypertension could affect placental function, thereby impacting the nutrient and hormone exchange critical for fetal development[19]. Hypertensive conditions might lead to altered levels of placental hormones, such as human chorionic gonadotropin, which is known to play a role in fetal thyroid function[20]. Additionally, the stress and inflammation associated with hypertension could lead to epigenetic modifications that affect gene expression related to thyroid development and function[21]. Moreover, maternal hypertension has been associated with oxidative stress and vascular dysfunction, creating a suboptimal in-utero environment that disrupts the endocrine axis and fetal thyroid gland maturation, contributing to congenital hypothyroidism[22, 23]. Understanding these pathways opens new avenues for research and highlights the need for careful monitoring and management of hypertensive conditions during pregnancy to mitigate potential risks to offspring health.

The finding that father’s diabetes is significantly associated with an increased risk of congenital malformations of male genital organs aligns with previous research. Ding’s study highlights how diabetes can impair male fertility by disrupting glucose metabolism, leading to poor sperm quality and DNA integrity. Diabetes can cause long-term damage and dysfunction in various organs, including the male reproductive system, affecting sperm motility and fertilization capacity[24]. Additionally, T. Fullston’s research suggests that father’s obesity can impair reproductive health across generations through epigenetic inheritance[25], while Michelle Lane’s review emphasizes the significant influence of both mother’s and father’s obesity on offspring health[26]. These findings underscore the importance of father’s preconception health, particularly concerning metabolic conditions, in influencing the risk of congenital anomalies in offspring.

The distinct causal relationships identified in this study highlight the need for targeted preconception and prenatal care strategies to manage parental health conditions, thereby potentially reducing the risk of congenital anomalies in offspring. Hypertension and diabetes, in particular, appear to play significant roles in the development of certain congenital anomalies, suggesting that genetic and possibly epigenetic mechanisms are involved. These findings call for further research to explore the biological pathways and to develop targeted interventions.

Future research should focus on validating these findings in diverse populations and investigating other potential parental health conditions. Additionally, studies should aim to identify biomarkers that can help stratify the risk of congenital anomalies based on parental health status. Clinical trials evaluating the efficacy and safety of interventions designed to manage these health conditions before and during pregnancy are essential.

This study has several strengths, including the use of robust MR techniques to minimize confounding and ensure reliable causal inferences. The comprehensive analysis using GWAS data from European ancestry populations minimized population stratification bias, and various sensitivity analyses confirmed the robustness of our findings. Furthermore, consistent results across different datasets underscored the robustness of our findings[27].

Our study has limitations affecting generalizability and interpretation. Focusing on a single ethnic group restricts broader applicability. Replication in diverse cohorts is necessary. Reliance on summary statistics from GWAS may introduce misclassification bias. Additionally, potential pleiotropy in Mendelian randomization analysis and residual confounding remains concerns, despite sensitivity analyses. Additionally, our MR analyses yielded extreme odds ratios (ORs), which could be attributed to the small number of instrumental variables or variability in the effect sizes of the genetic instruments used. These extreme values should be interpreted with caution, and further analyses are needed to confirm these findings.

**In conclusion**, our study offers compelling evidence of significant causal relationships between certain parental illnesses and heightened risk of congenital anomalies. The insight holds promise for improving clinical outcomes among offspring at risk of congenital anomalies.

## Data Availability

All data produced in the present study are available upon reasonable request to the authors.

https://gwas.mrcieu.ac.uk/

## Acknowledgments

We want to acknowledge the participants and investigators of the FinnGen study, UK Biobank, and other GWASs included in this study. Without the dedication of these organizations and their members, this article would have been difficult to complete.

## Clinical trial number

Not applicable

## Ethics Statement

Not applicable

## Author Contributions

WJ.S. conceived the project, designed the study and revised the manuscript.; T.T. wrote the manuscript; L.T. collected data from the public database, and all authors reviewed the manuscript.

## Consent for Publication

Not applicable

## Data Availability Statement

The original contributions presented in the study are included in the article/Supplementary Material. Further inquiries can be directed to the corresponding author.

## Abbreviations

CA: Congenital anomalies
CM: congenital malformations
CD: congenital deformities
IVW: Inverse variance weighted
IVs: Instrumental variables
SNPs: Single Nucleotide Polymorphisms
MR: Mendelian randomization
UVMR: Univariable MR
MVMR: Multivariable MR
OR: Odds ratio
CI: confidence interval
GWAS: Genome-Wide Association Study
UKBB: United Kingdom Biobank

