## Supplementary figures for "Mendelian randomization infers the effect of 14 parental illnesses on 44 congenital anomalies"

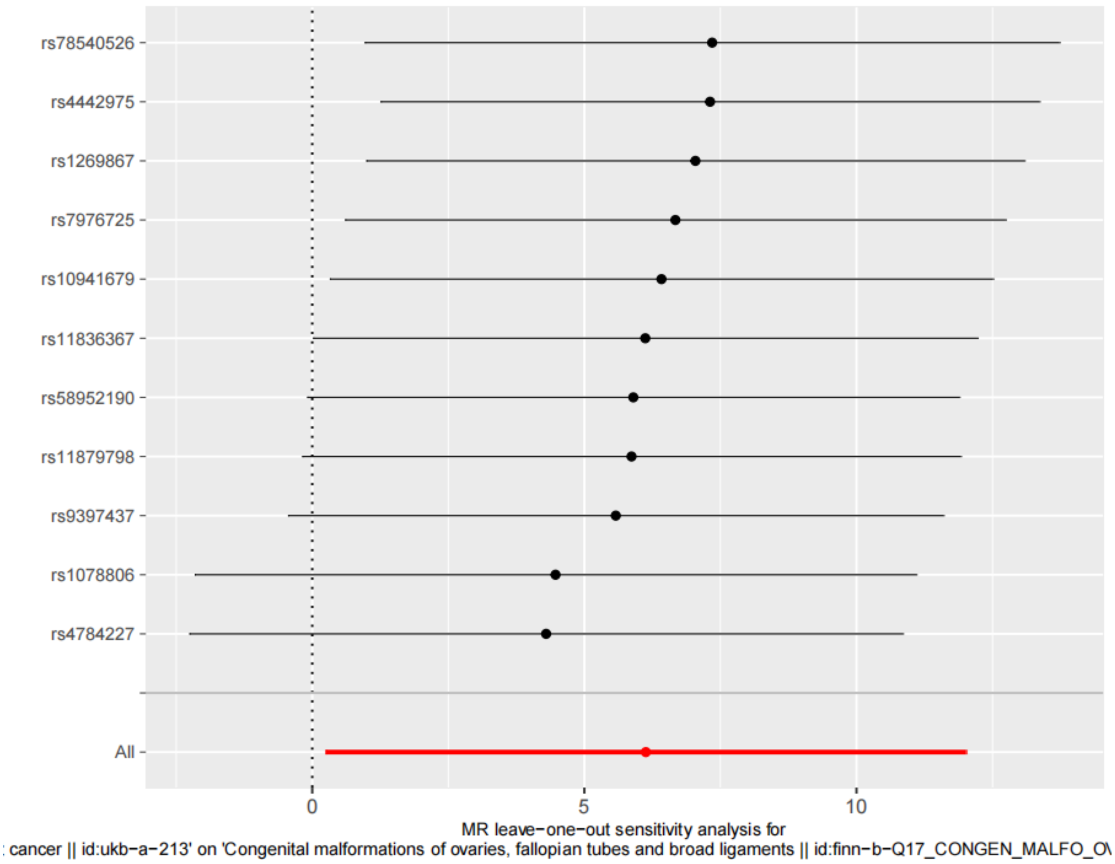


Supplementary Figures 1. Leave-one-out sensitivity analysis plots for Illnesses of mother: breast cancer on CM of the ovaries, fallopian tubes, and broad ligaments. CA, congenital anomalies; CD, congenital deformities; CM, congenital malformations; MR, mendelian randomization.

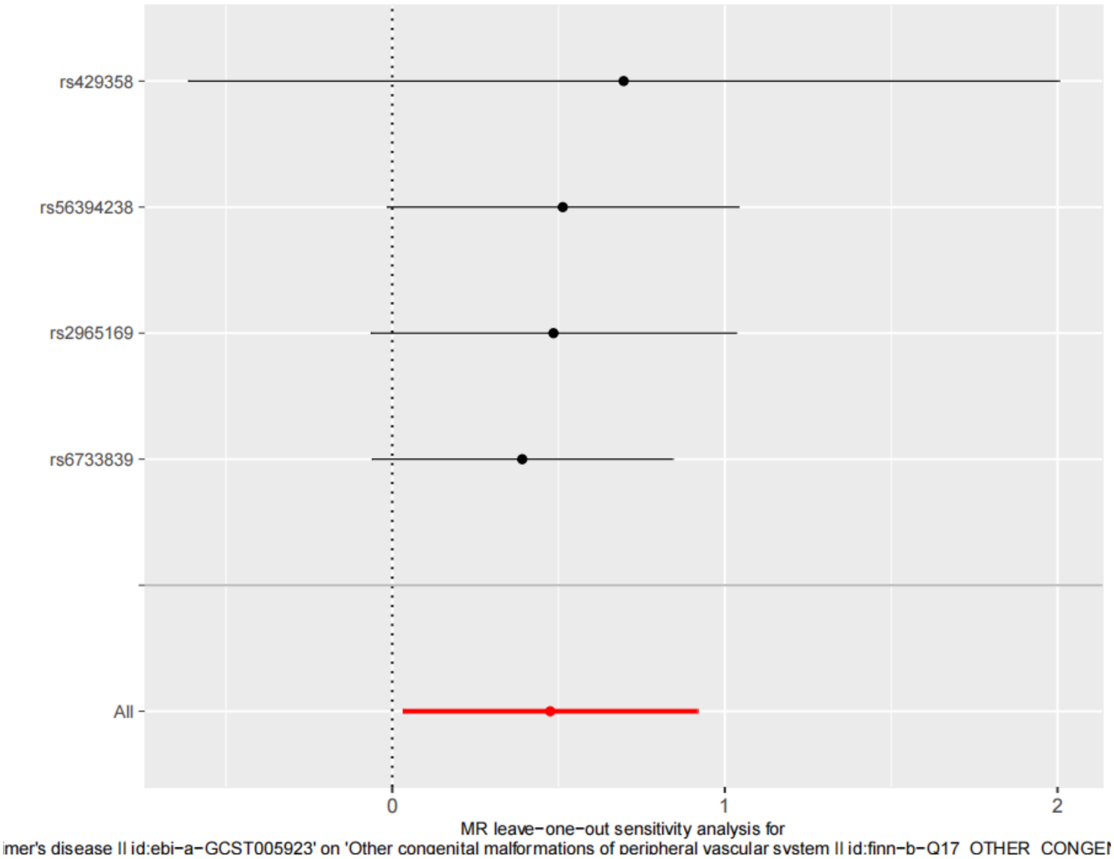


Supplementary Figures 2. Leave-one-out sensitivity analysis plots for Illnesses of mother: Alzheimer's disease with other CM of the peripheral vascular system. CA, congenital anomalies; CD, congenital deformities; CM, congenital malformations; MR, mendelian randomization.

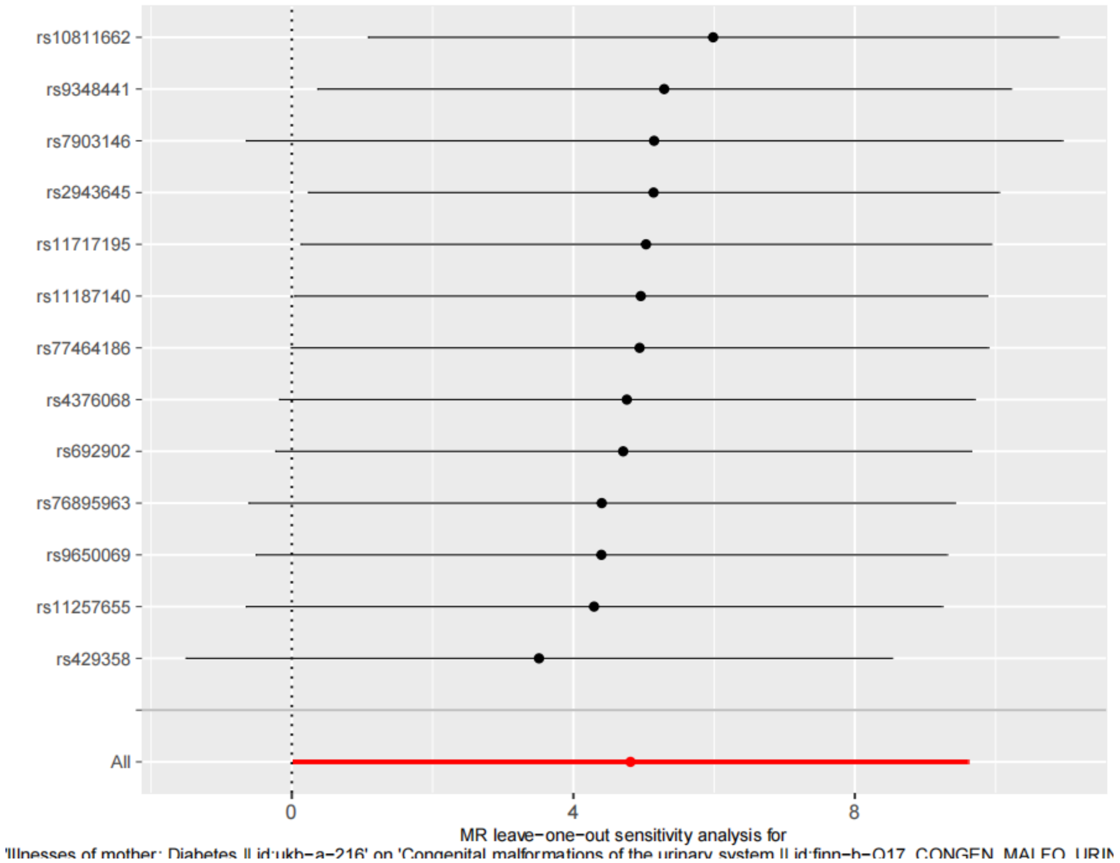


Supplementary Figures 3. Leave-one-out sensitivity analysis plots for Illnesses of mother: diabetes with CM of the urinary system. CA, congenital anomalies; CD, congenital deformities; CM, congenital malformations; MR, mendelian randomization.

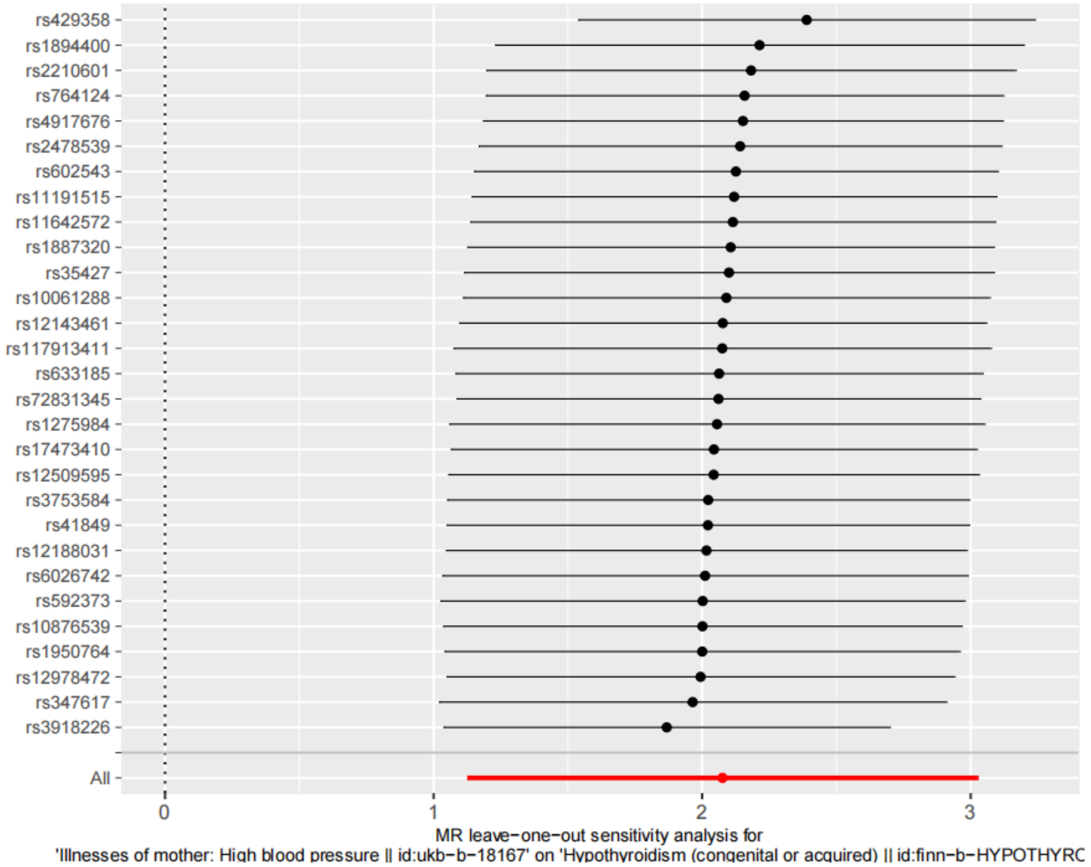


Supplementary Figures 4. Leave-one-out sensitivity analysis plots for Illnesses of mother: hypertension with congenital hypothyroidism. CA, congenital anomalies; CD, congenital deformities; CM, congenital malformations; MR, mendelian randomization.

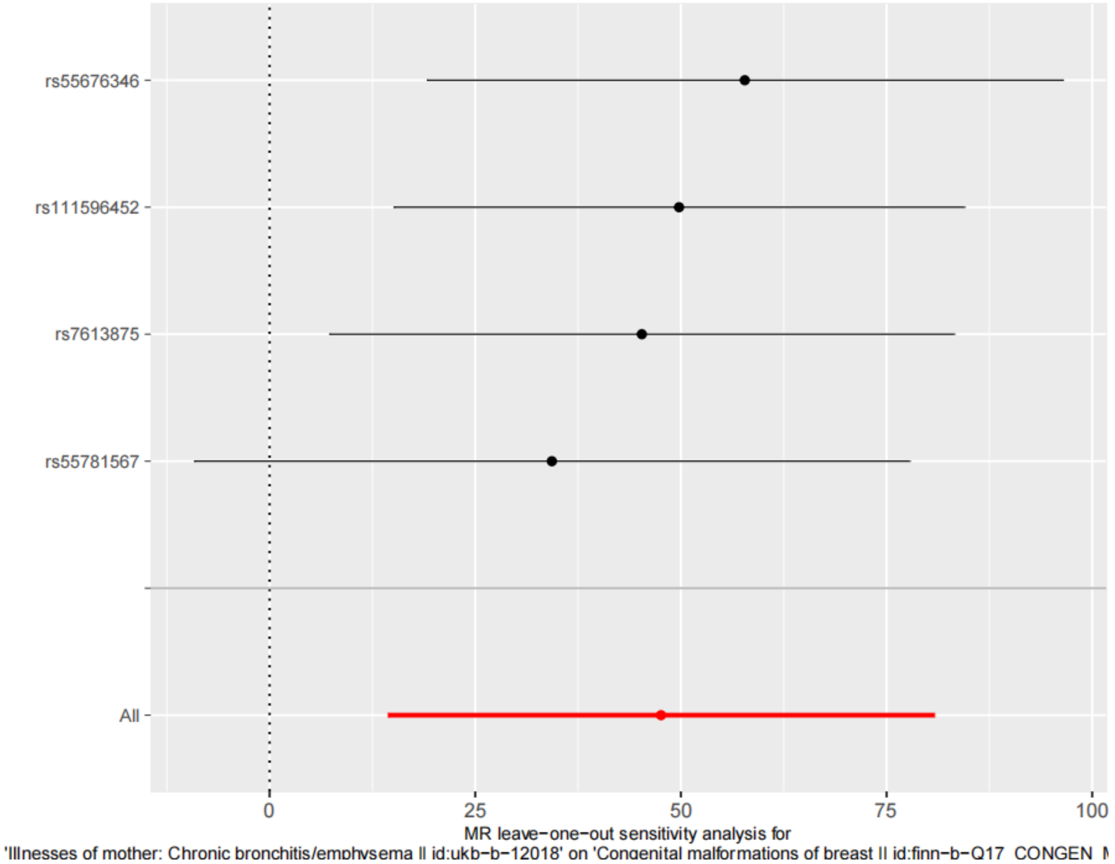


Supplementary Figures 5. Leave-one-out sensitivity analysis plots for Illnesses of mother: chronic bronchitis/emphysema with CM of the breast. CA, congenital anomalies; CD, congenital deformities; CM, congenital malformations; MR, mendelian randomization.

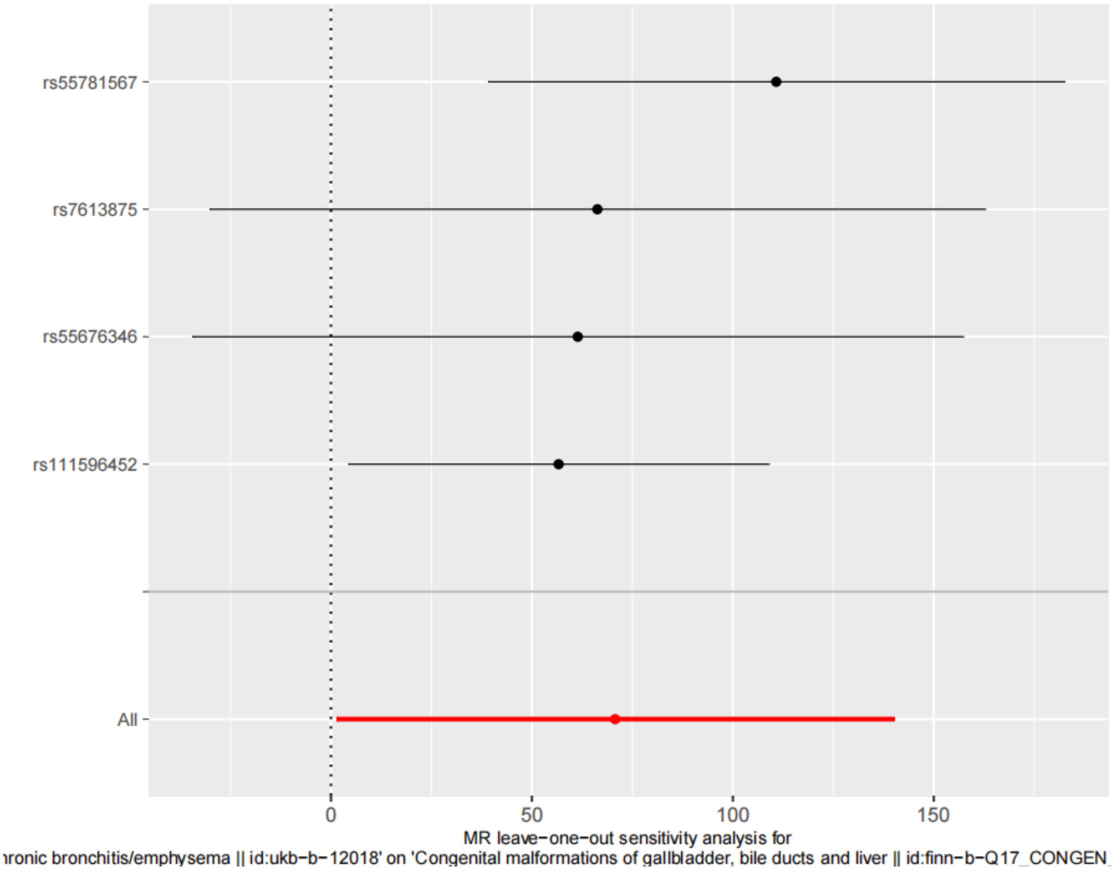


Supplementary Figures 6. Leave-one-out sensitivity analysis plots for Illnesses of mother: chronic bronchitis/emphysema with CM of the gallbladder, bile ducts, and liver. CA, congenital anomalies; CD, congenital deformities; CM, congenital malformations; MR, mendelian randomization.

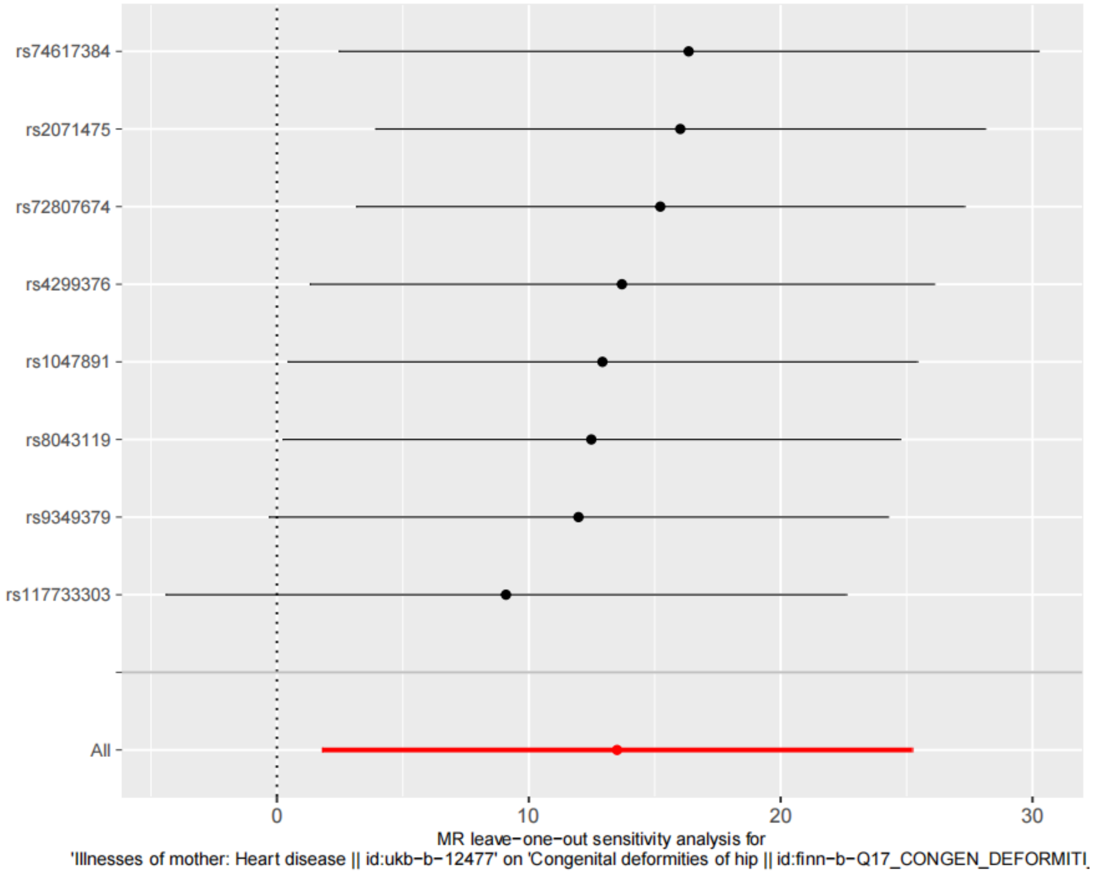


Supplementary Figures 7. Leave-one-out sensitivity analysis plots for Illnesses of mother: heart disease with congenital deformities (CD) of the hip. CA, congenital anomalies; CD, congenital deformities; CM, congenital malformations; MR, mendelian randomization.

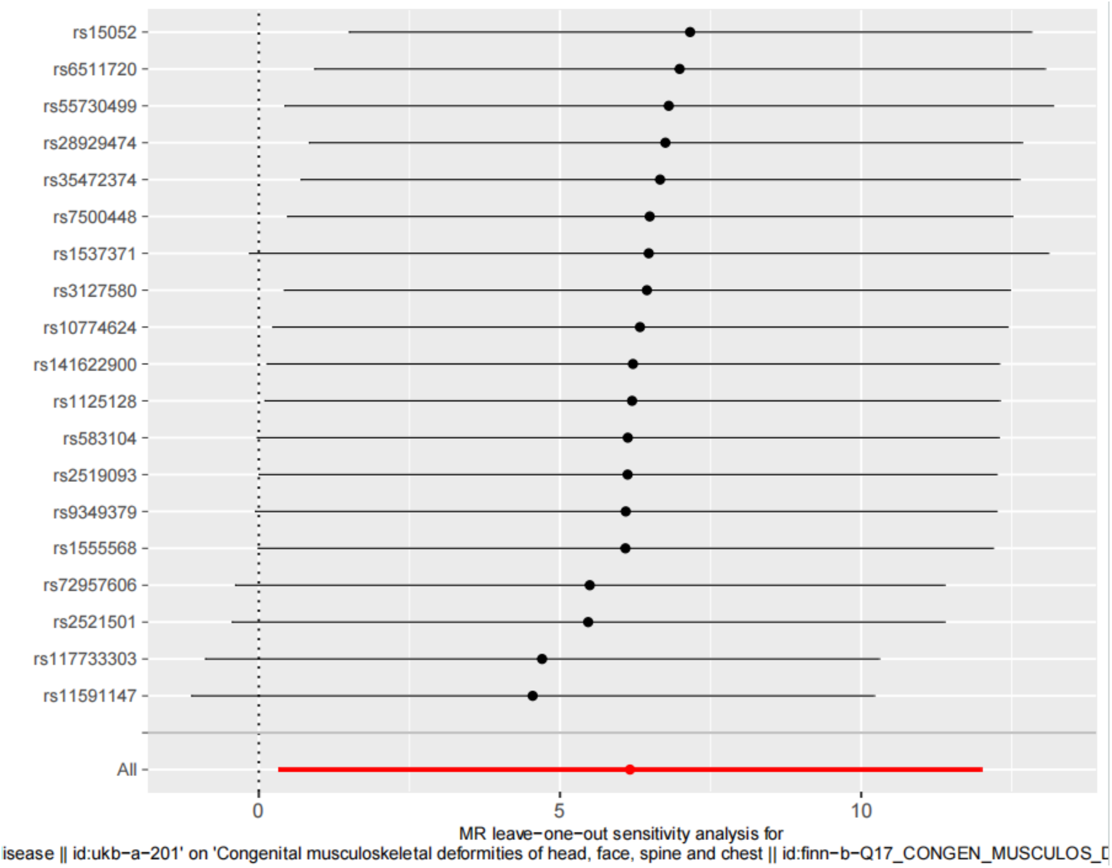


Supplementary Figures 8. Leave-one-out sensitivity analysis plots for Illnesses of father: heart disease with CD of the head, face, spine, and chest. CA, congenital anomalies; CD, congenital deformities; CM, congenital malformations; MR, mendelian randomization.

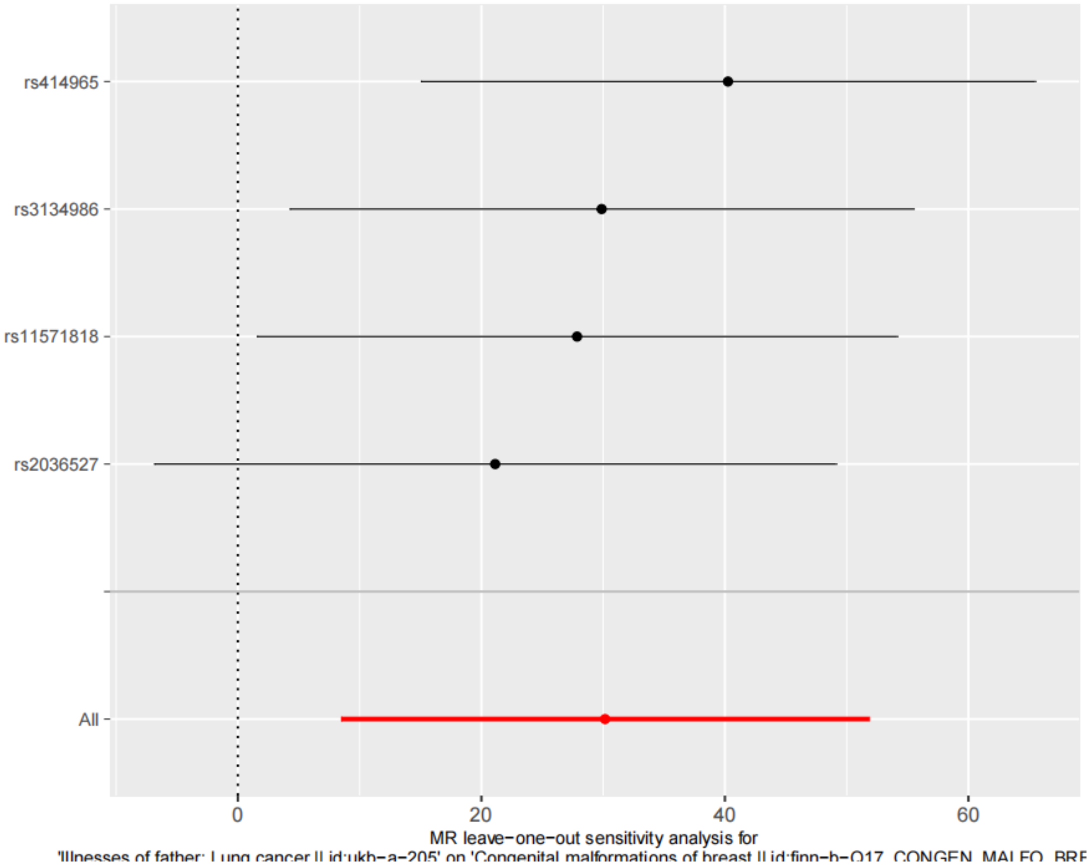


Supplementary Figures 9. Leave-one-out sensitivity analysis plots for Illnesses of father: lung cancer with CM of the breast. CA, congenital anomalies; CD, congenital deformities; CM, congenital malformations; MR, mendelian randomization.

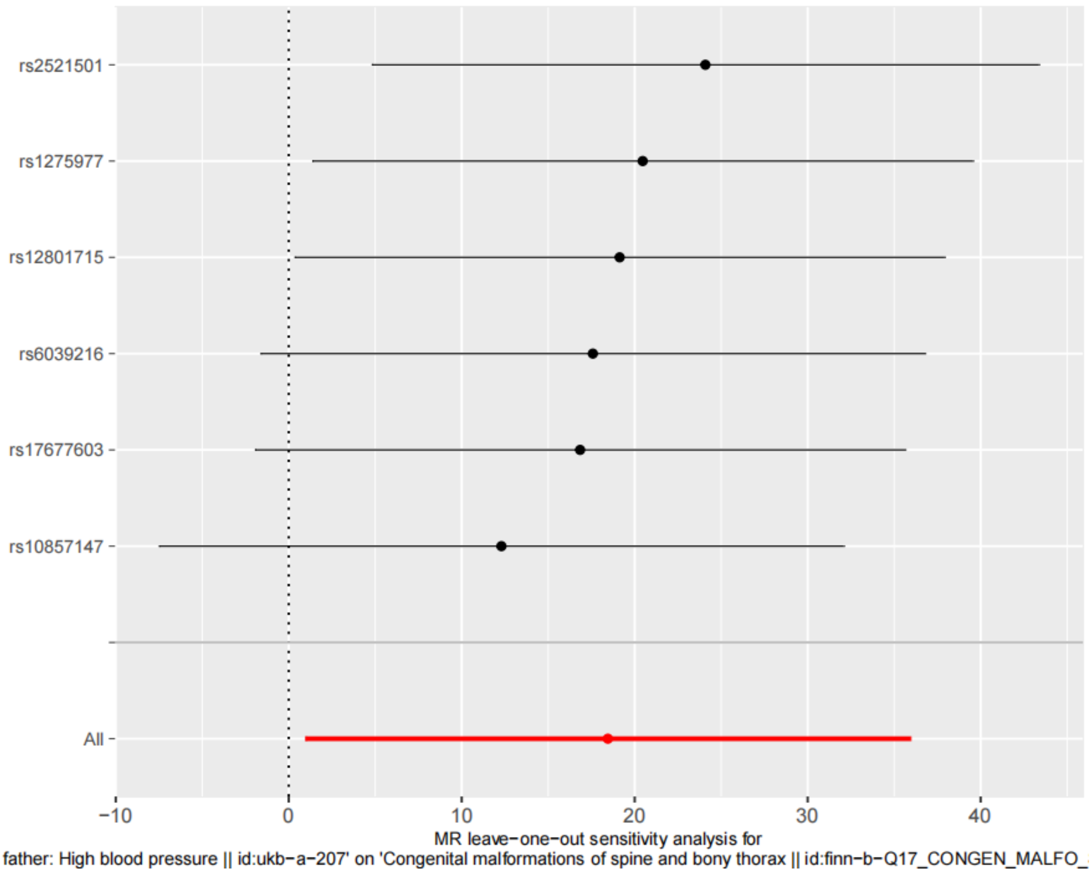


Supplementary Figures 10. Leave-one-out sensitivity analysis plots for Illnesses of father: hypertension with CM of the spine and bony thorax. CA, congenital anomalies; CD, congenital deformities; CM, congenital malformations; MR, mendelian randomization.

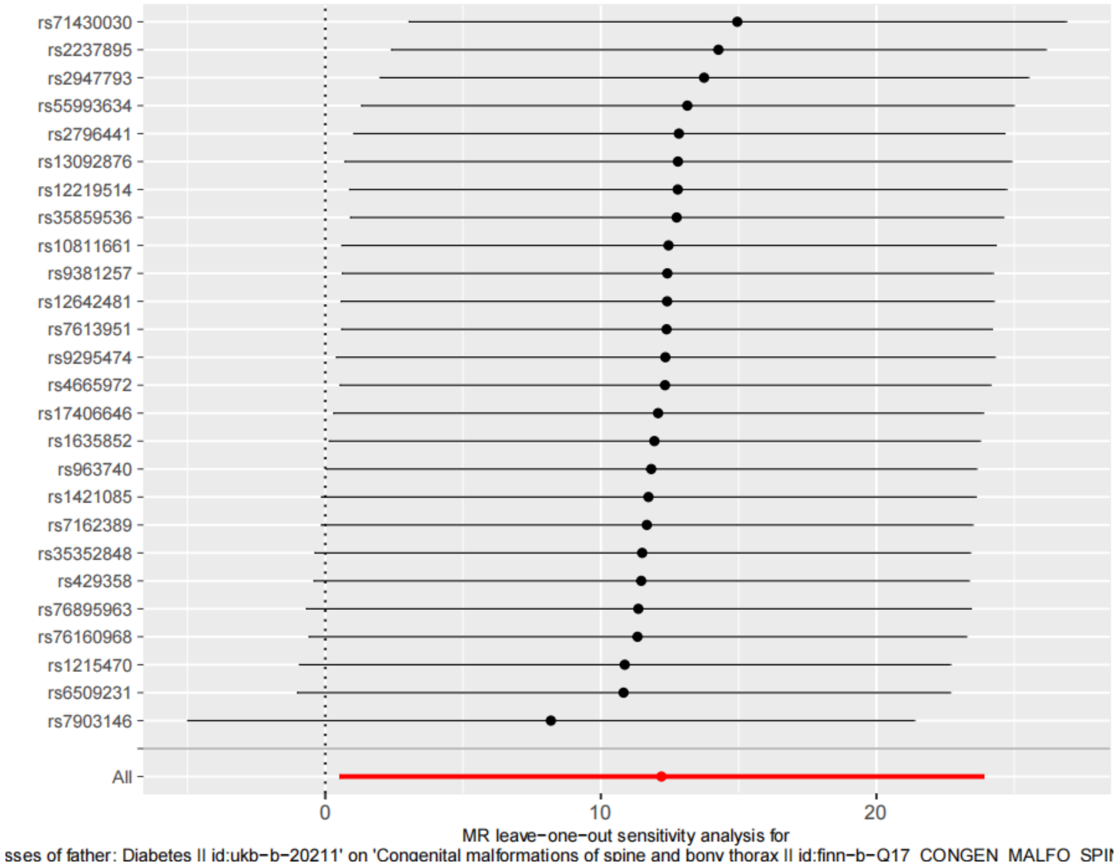


Supplementary Figures 11. Leave-one-out sensitivity analysis plots for Illnesses of father: diabetes with other CM of male genital organs. CA, congenital anomalies; CD, congenital deformities; CM, congenital malformations; MR, mendelian randomization.

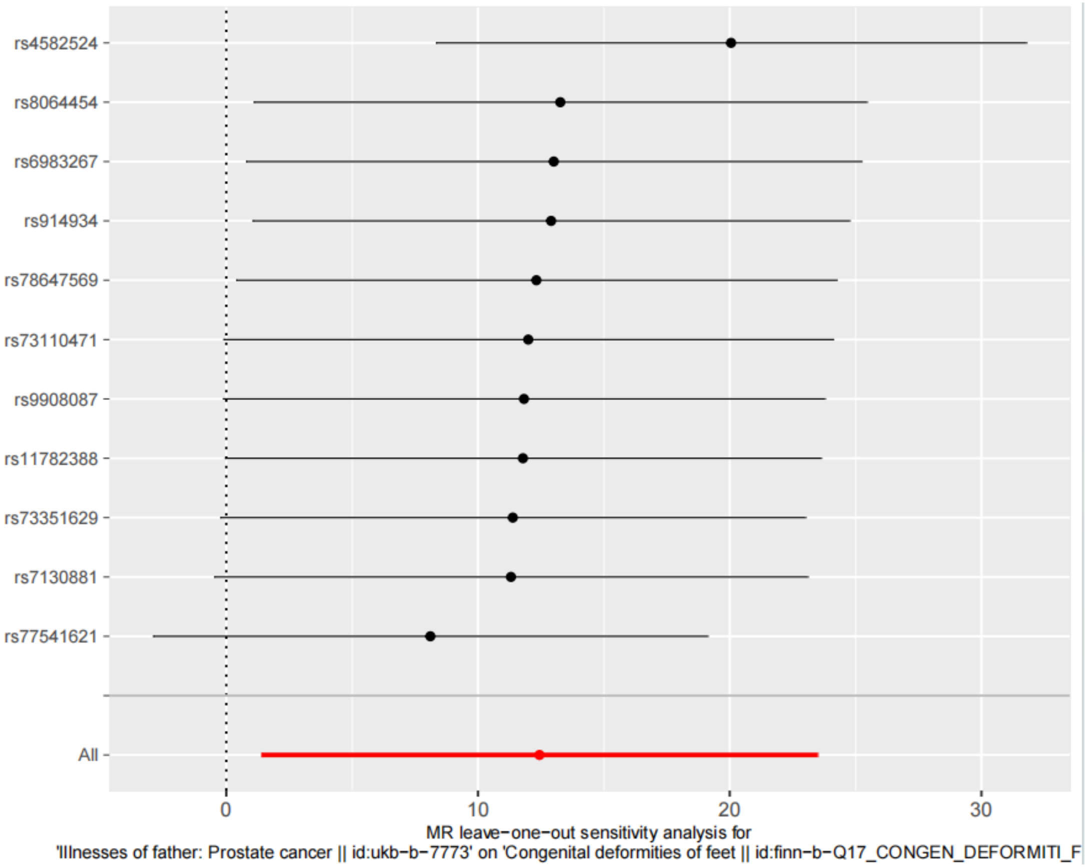


Supplementary Figures 12. Leave-one-out sensitivity analysis plots for Illnesses of father: prostate cancer with CD of the feet. CA, congenital anomalies; CD, congenital deformities; CM, congenital malformations; MR, mendelian randomization.

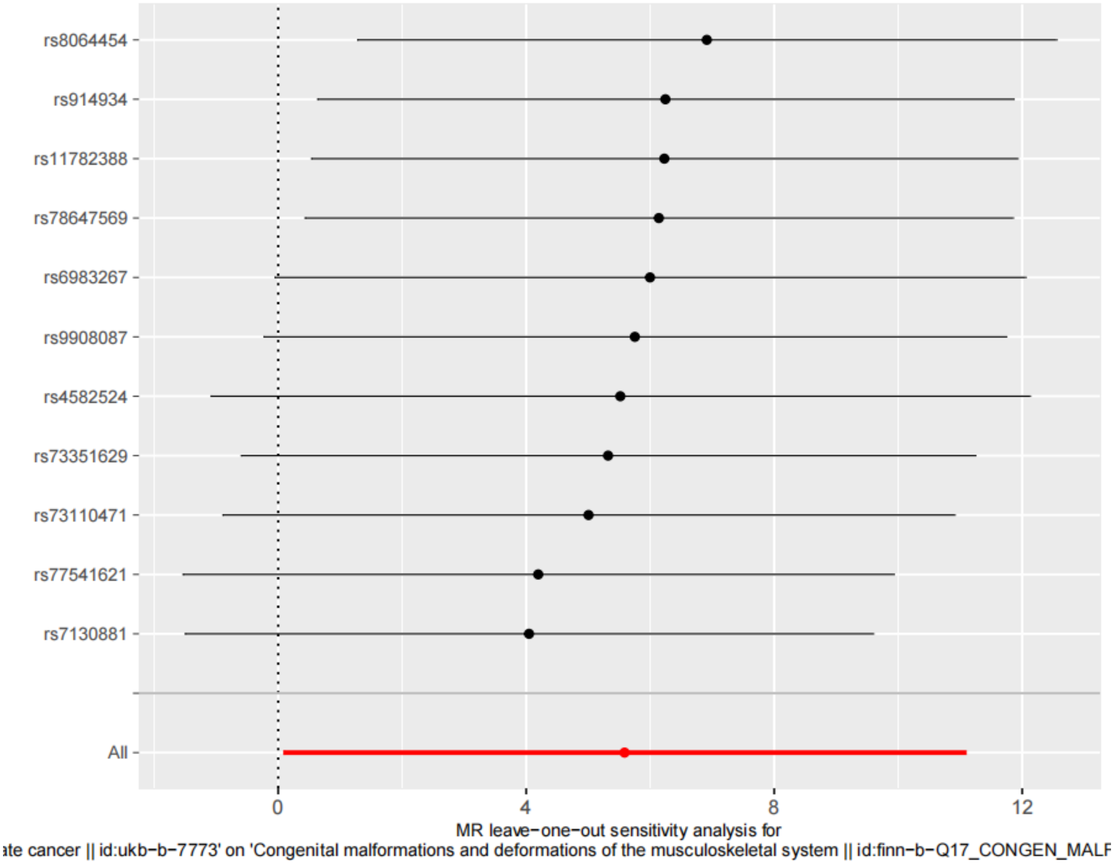


Supplementary Figures 13. Leave-one-out sensitivity analysis plots for Illnesses of father: prostate cancer with CM and CD of the musculoskeletal system. CA, congenital anomalies; CD, congenital deformities; CM, congenital malformations; MR, mendelian randomization.

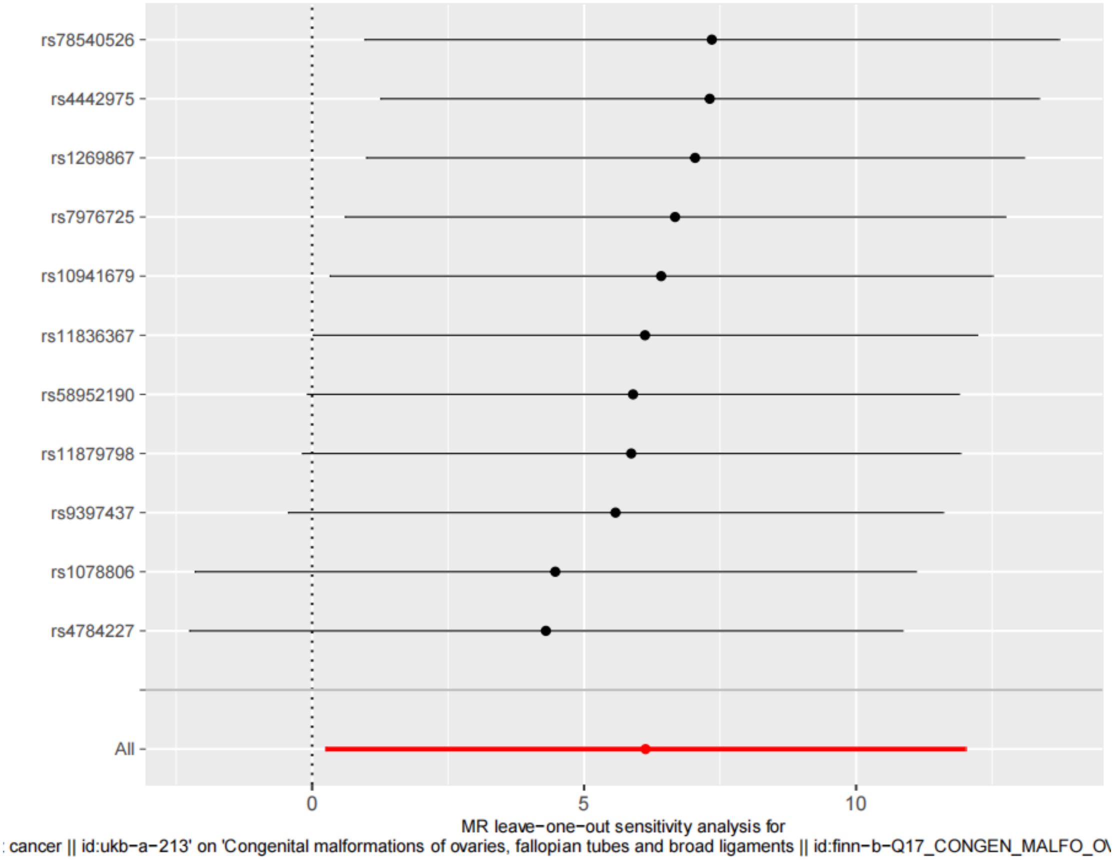


Supplementary Figures 1. Leave-one-out sensitivity analysis plots for breast cancer on CM of the ovaries, fallopian tubes, and broad ligaments. CA, congenital anomalies; CD, congenital deformities; CM, congenital malformations; MR, mendelian randomization.
